# COVID-19 vaccination and menstrual cycle changes: A United Kingdom (UK) retrospective case-control study

**DOI:** 10.1101/2021.11.23.21266709

**Authors:** Alexandra Alvergne, Gabriella Kountourides, M. Austin Argentieri, Lisa Agyen, Natalie Rogers, Dawn Knight, Gemma C Sharp, Jacqueline A Maybin, Zuzanna Olszewska

**Affiliations:** ISEM, Univ Montpellier, CNRS, IRD, Montpellier, France; School of Anthropology and Museum Ethnography, Oxford, UK; Harvard/MGH Center on Genomics, Vulnerable Populations, and Health Disparities, Massachusetts General Hospital, Boston, USA; Long Covid Support; MRC Integrative Epidemiology Unit, University of Bristol, UK; Bristol Medical School, Population Health Sciences, University of Bristol, UK; MRC Centre for Reproductive Health, University of Edinburgh, UK

## Abstract

**Background:** There has been increasing public concern that COVID-19 vaccines cause menstrual cycle disturbances, yet there is currently limited data to evaluate the impact of vaccination on menstrual health. Our objectives were (1) to evaluate the prevalence of menstrual changes following vaccination against COVID-19, (2) to test potential risk factors for any such changes, and (3) to identify patterns of symptoms in participants’ written accounts.

**Methods:** We performed a secondary analysis of a retrospective online survey titled “The Covid-19 Pandemic and Women’s Reproductive Health”, conducted in March 2021 in the UK before widespread media attention regarding potential impacts of SARS-CoV-2 vaccination on menstruation. Participants were recruited via a Facebook ad campaign in the UK and eligibility criteria for survey completion were age greater than 18 years, having ever menstruated and currently living in the UK. In total, 26,710 people gave consent and completed the survey. For this analysis we selected 4,989 participants who were pre-menopausal and vaccinated. These participants were aged 28 to 43, predominantly from England (81%), of white background (95%) and not using hormonal contraception (58%).

**Findings:** Among pre-menopausal vaccinated individuals (n=4,989), 80% did not report any menstrual cycle changes up to 4 months after their first COVID-19 vaccine injection. Current use of combined oral contraceptives was associated with lower odds of reporting any changes by 48% (OR = 0.52, 95CI = [0.34 to 0.78], *P*<0.001). Odds of reporting any menstrual changes were increased by 44% for current smokers (OR = 1.44, 95CI = [1.07 to 1.94], *P*<0.01) and by more than 50% for individuals with a positive COVID status [Long Covid (OR = 1.61, 95CI = [1.28 to 2.02], *P*<0.001), acute COVID (OR = 1.54, 95CI = [1.27 to 1.86], *P*<0.001)]. The effects remain after adjusting for self-reported magnitude of menstrual cycle changes over the year preceding the survey. Written accounts report diverse symptoms; the most common words include “cramps”, “late”, “early”, “spotting”, “heavy” and “irregular”, with a low level of clustering among them.

**Conclusions:** Following vaccination for COVID-19, menstrual disturbance occurred in 20% of individuals in a UK sample. Out of 33 variables investigated, smoking and a previous history of SARS-CoV-2 infection were found to be risk factors while using oestradiol-containing contraceptives was found to be a protective factor. Diverse experiences were reported, from menstrual bleeding cessation to heavy menstrual bleeding.

## Introduction

There has been increasing public concern that COVID-19 vaccines cause disruption of menstrual cycles [1–3], leading to problematic menstrual symptoms, vaccine hesitancy [4] and fears about the impact of vaccination on fertility [5–7]. There are currently limited data [8] for investigating the relationship between the COVID-19 vaccines and menstrual cycles [1,9,10]. This is despite rising awareness among clinicians that the menstrual cycle should be used as a vital sign of female health [11,12], that sex is a biological variable which should be considered in immunological studies [13] and that there have been reports of heavy, infrequent or irregular menstrual bleeding following vaccination [1,8–10]. Quantitative evidence for any such relationship between COVID-19 vaccination and menstrual cycle disturbance, as well as the factors mediating this relationship, are crucial for evaluating how female health has been impacted by the pandemic.

The first published study on the topic of vaccine effects on menstrual cycles dates back to 1913, when a medical doctor at the Presbyterian Hospital, New York, concluded that there was a striking relationship between the prophylactic typhoid vaccine and menstrual disturbances among one hundred cases [14]. After ruling out all other apparent causes, he found that 53% showed some type of disturbance, including increased or decreased frequency, increased or decreased volume and dysmenorrhoea [14]. These disturbances disappeared within 6 months of the vaccine, suggesting that any such vaccine side-effect was temporary. There has also been a report of menstrual disturbances following inoculation with the hepatitis vaccine in a Japanese study conducted in 1982. Among 16 hospital employees, 7 reported various menstrual abnormalities including decreased volume of menstruation, infrequent or too frequent menses [15]. The changes were attributed to the use of human plasma to make the vaccine (antigens were derived from human plasma, containing hormonal impurities). More recently, large-scale studies on the effects of vaccination on menstrual disturbances reported mixed results. A 2018 study of 29,846 female residents of Nagoya City, Japan, found that none of the 24 symptoms investigated, including menstrual symptoms, were associated with increased odds of occurring after administration of the HPV vaccine. However, age-adjusted odds of hospital visits were increased for “abnormal amount of menstrual bleeding” (OR=1.43, 95%CI=[1.13 to 1.82]), “irregular menstruation” (OR=1.29, 95%CI=[1.12 to 1.49]) and chronic, persisting “abnormal amount of menstrual bleeding” (OR 1.41, 95% CI: 1.11–1.79)[16]. Although retrospective and sensitive to recall bias among those receiving the vaccine, the study suggests a possible link between the HPV vaccine and menstrual irregularities. Another study applying a signal detection analysis on the FDA Vaccine Adverse Event Reporting System (VAERS) shows a disproportionate number of reports of premature ovarian insufficiency, amenorrhea, irregular menstruation, increase in FSH and premature menopause following administration of the HPV vaccine [17]. However, the evidence is non-causal, and relationships might depend on the type of vaccine. With regards to COVID-19, the UK’s Medicine and Healthcare products Regulatory Agency (MHRA) is closely monitoring reports of menstrual disorders [18], with more than 30,000 reports made to its yellow card surveillance scheme by 2 September 2021 for both mRNA and adenovirus-vectored COVID-19 vaccines [19]. Recent data from a gender-diverse sample receiving COVID-19 vaccination in the US suggests that changes in the form of heavy and breakthrough bleeding affect many people. However, there has been no quantitative assessment of the risk factors for menstrual disturbances following COVID-19 vaccination prior to widespread media attention ([8], Box 1).

### Objectives of the study

The objectives of this study are three-fold: (1) to evaluate the incidence of reports of menstrual changes of any kind following COVID-19 vaccination in a sample broadly representative of those who menstruate in the UK, (2) to investigate the risk factors for reporting any menstrual changes following COVID-19 vaccination, and (3) to capture the types and breadth of menstrual disturbances by analysing the text written by participants. We build on a large retrospective cross-sectional study on menstruation during the pandemic conducted in the UK, launched before UK media coverage of concerns over menstrual vaccine side-effects and including both quantitative and textual data on menstrual cycle changes perceived to be induced by the COVID-19 vaccines.

## Methods

### Study design

The online survey was initially designed to evaluate whether and how the COVID-19 pandemic influenced menstrual health. Retrospective and self-reported data on menstrual cycles, behaviour, life circumstances and health before and during the pandemic as well as SARS-CoV-2 infection and vaccination status were collected using an online survey hosted on the Qualtrics platform (www.qualtrics.com). All survey responses were anonymized using randomly generated IDs. The study, titled “The Covid-19 Pandemic and Women’s Reproductive Health” has been reviewed by, and received ethics clearance through, the Oxford University School of Anthropology and Museum Ethnography Departmental Research Ethics Committee [SAME_C1A_20_029].

### Patient and Public Involvement

During the design of survey questions, input from a panel of women suffering from Long Covid, referred to us by the Long Covid Support online group (https://www.longcovid.org/), was incorporated. The results were discussed with panel members who were also invited to co-author the paper and co-design dissemination plans.

### Study population

The online survey was launched on March 8, 2021. The title of the survey was kept general (“female reproductive health and the COVID pandemic”) so as not to oversample individuals with specific interest in menstrual cycles and COVID infection or vaccination. The survey was disseminated through a Facebook advertising campaign, and included images of women of diverse ethnicities, ages, and abilities, as well as images of breastfeeding and pregnant women (SI1); we fine-tuned the ad targeting (to the extent that Facebook allows) throughout the campaign to ensure even geographical and socio-economic spread. As explained in the information page (SI2), participants could only complete the survey if they were over 18, had ever menstruated, currently lived in the UK, and gave informed consent to the use of their data. The survey included a maximum of 105 questions depending on individual circumstances (SI3) and took an average of 24 minutes to complete. Of the eligible participants who started the survey, 61% answered all questions after giving their consent (on average participants completed 80% of the questionnaire). In case of survey fatigue, progress could be saved for up to 14 days to allow participants to resume later. The survey was disseminated through a Facebook advertisement campaign targeting all menstruators in the UK, from 08/03/21 to 01/06/21, at which point there had been no new entries for a week. During the campaign, we used a stratified sampling strategy to ensure that subgroups of the UK population in terms of age, income and ethnicity were represented in the final sample. In total, 695,543 people viewed the survey ad on their Facebook page and 26,710 with eligible criteria gave consent and completed it (there were no duplicates), leading to a 3.8% response rate. The data, data dictionary and scripts are available on the Open Science Framework Platform (https://osf.io/pqxy2/).

### Outcome: vaccine side-effects on menstrual cycles

While the survey did not initially aim to evaluate the impact of vaccination on menstrual cycles specifically, a question was included to assess participants’ perception of their menstrual cycles following vaccination at the end of the survey. Specifically, participants who indicated that they had been menstruating in the past 12 months, received 1 or 2 doses of the COVID-19 vaccines and were not involved in a clinical trial were asked “*Have you noticed any changes to your menstrual cycles since you got vaccinated?*”, to which 1 of 4 possible answers could be given: “No”, “Yes, my menstrual cycles are MORE disrupted”, “Yes, my menstrual cycles are LESS disrupted”, “Other (please state)”. Although “disruption” per se was not defined, by the time participants answered this question, they had already completed many questions on menstrual cycle regularity, duration, and symptoms. At the time of the survey design, anecdotal reports of menstrual effects of the vaccine were only just beginning to circulate, while people with Long Covid were reporting either improvement or worsening of their symptoms in general after vaccination. This question was included with the intention of investigating the latter effects. Participants could select the answer “Other”, which in some cases may not have been a different decision from choosing either “more disrupted” or “less disrupted”. For analysis, we thus transformed these variables to represent a binary outcome (“No changes” vs. “Any other changes”).

### Exposures

A total of 33 variables were extracted for this analysis. In addition to socio-demographic variables (age, income, education, gender, ethnic group, marital status), and standard proxies for health (BMI, smoking status, physical activity, regular use of vitamins/supplements, regular use of medicine), the dataset included vaccine-related, COVID and pandemic-related, and reproductive variables (See SI4 for the operationalization of variables). First, data on the type of vaccine received, of which only two had been approved for use in the UK at the time (Pfizer BioNTech/Oxford-AstraZeneca/Not sure), and the timing of the first vaccination (month/year) were included. Second, COVID status was operationalized in two ways: (i) based on whether people thought they had had COVID, as widespread testing had not been available in the UK in the early months of the pandemic which fell within the survey period, leading to three categories: *No COVID, acute COVID* (symptoms lasting less than 28 days) and *Long Covid* (symptoms lasting more than 28 days) as well as (ii) based on a combination of testing and self-diagnosis, leading to three categories: *No COVID* (no tests or negative tests), *COVID tested +* (positive test) and “*Self-diagnosed positive”* (referring to individuals who had a suspected or clinically diagnosed COVID infection but had not obtained positive PCR, antigen or antibody tests). We included this last category due to the unavailability of widespread testing in the UK in the first wave of the pandemic in 2020 and ongoing questions about the accuracy and optimal timing of antigen and antibody tests. In addition, variables indicative of changes in both life satisfaction and menstrual cycle symptoms compared to before the pandemic were also included to adjust for changes experienced because of the pandemic and/or the infection rather than vaccination. Third, reproductive variables indicative of menstrual health before the pandemic (age at menarche, cycle length, period length, cycle irregularity, heavy bleeding), reproductive history (number of deliveries) and contraceptive use were included.

### Statistical analysis

The aim of the quantitative analysis was two-fold: (1) to quantify the extent to which individuals answered “No changes” when asked about any perceived changes to their menstrual cycle following COVID-19 vaccination, and (2) to evaluate potential risk and protective factors for selecting any other answer. The original outcome variable is nominal (two or more categories with no intrinsic order) but violates the IIA assumption (Independence or Irrelevant Alternatives) as options were not independent, thus we dichotomized the variable into two mutually exclusive categories (“No changes”, “Any other changes”) and performed logistic regressions. We first conducted a series of exploratory univariable analyses, investigating each of 33 variables as potential risk factors for reporting changes in menstrual cycles following vaccination. We then retained all variables significant at the false discovery rate (FDR) threshold (FDR-corrected *P*<0.05) [20] for consideration in multivariable analyses. We then conducted separate multivariable analyses with each of the variables identified in the univariable analyses as exposures variables. Each multivariable model was adjusted for potential confounders, which were defined as variables significant at the FDR threshold in the univariable analyses and with a potential confounding (but not mediating) effect according to hypothesized directed acyclic graphs (DAG, SI5). Estimates and confidence intervals on the log-odds scale were converted to odds-ratios for reporting. To test the significance of individual coefficients, p-values were derived from Wald χ^2^ statistics. For all models, we plotted a receiver operating characteristic curve (ROC) and computed a measure of the accuracy of the chosen model in predicting the outcome using the area under the curve (AUC). As an alternative way of selecting covariates for the multivariable models, and to improve model prediction accuracy, we also performed LASSO regression using the “*glmnet*” package in R [21]. As the range and scale of variables can influence the penalization for having too many variables in elastic net models, all ordinal variables were coded numerically and re-classed as continuous, and all continuous variables were centered and standardized. Nominal categorical variables were broken out into individual binary dummy variables for all response levels except for the reference level.

### Missing data

The analysis of complete cases only can introduce bias and lead to a substantial reduction of statistical power [22], especially if it is plausible that the data are missing at random or not completely at random. An evaluation of the missing data suggested that multiple imputation was advisable (SI6). The average proportion of missing values across all variables in the dataset was 3.8%, which was mostly accounted for by the variable BMI (38% of missing data, SI6). To handle missing data, we used a multiple imputation approach using the R package ‘*missRanger’* [23], which combines random forest imputation with predictive mean matching [23]. Prior to all analyses, we imputed 5 datasets, with a maximum of 10 iterations specified for each imputation. Each imputation was also weighted by the degree of missing data for each participant, such that the contribution of data from participants with higher proportions of missingness was weighted down in the imputation. We set the maximum number of trees for the random forest to 200 but left all other random forest hyperparameters at their default. The average out-of-bag (OOB) error rate for multiple imputation across all imputed datasets was 0.08 in women (range: 0 to 0.77) and 0.08 in men (range: 0 to 0.69). Parameter estimates for all five datasets were pooled to provide more accurate estimates. A sensitivity analysis was also performed on the complete cases without missing data imputation (n=1,548 (SI7)).

### Text analysis

We first built a custom text cleaning function using the *‘textclean’* [24] and ‘*tidytext’* [25] R packages to analyse the text written by participants selecting the “Other” category in the outcome variable (n=574). The resulting corpus was tokenized (broken into individual units) and lemmatized (words derived from others, such as “vaccine” and “vaccination” were grouped by their stem version “vaccine” (SI8). The corpus was analysed to answer the following 3 questions: (i) which single words (unigrams) and pairs of adjacent words (bigrams) are most frequent? (ii) which words co-occur in the same sentence? (iii) Are there clusters of symptoms? To investigate the commonality of words, we explored the frequency of unigrams and bigrams within all responses. We performed a correlation analysis on the most important words for menstrual cycle descriptions to measure the association between words using the correlation index (phi coefficient (φ)). To explore patterns of symptoms we examined the words that commonly occur together (though not necessarily adjacent) to visualize groups of words that cluster together. Clusters were visualized by arranging correlated words into a combination of connected nodes (network graph) using the ‘*igraph’* package [26].

## Results

Out of the 26,710 individuals who completed the survey, 8,539 (32%) reported having been vaccinated, with either 1 (n=7,270) or 2 doses (n=1,269). In the final sample, we only included individuals living in the UK who knew about their vaccination status, who had a period in the last 12 months and who were also pre-menopausal and not pregnant. We also excluded participants who selected “Other changes” and contributed text to the effect of “too early to say” when describing menstrual disturbances following COVID-19 vaccination (n=369, 64% of those selecting the answer “Other changes)” (Fig. 1)

**Figure 1.**
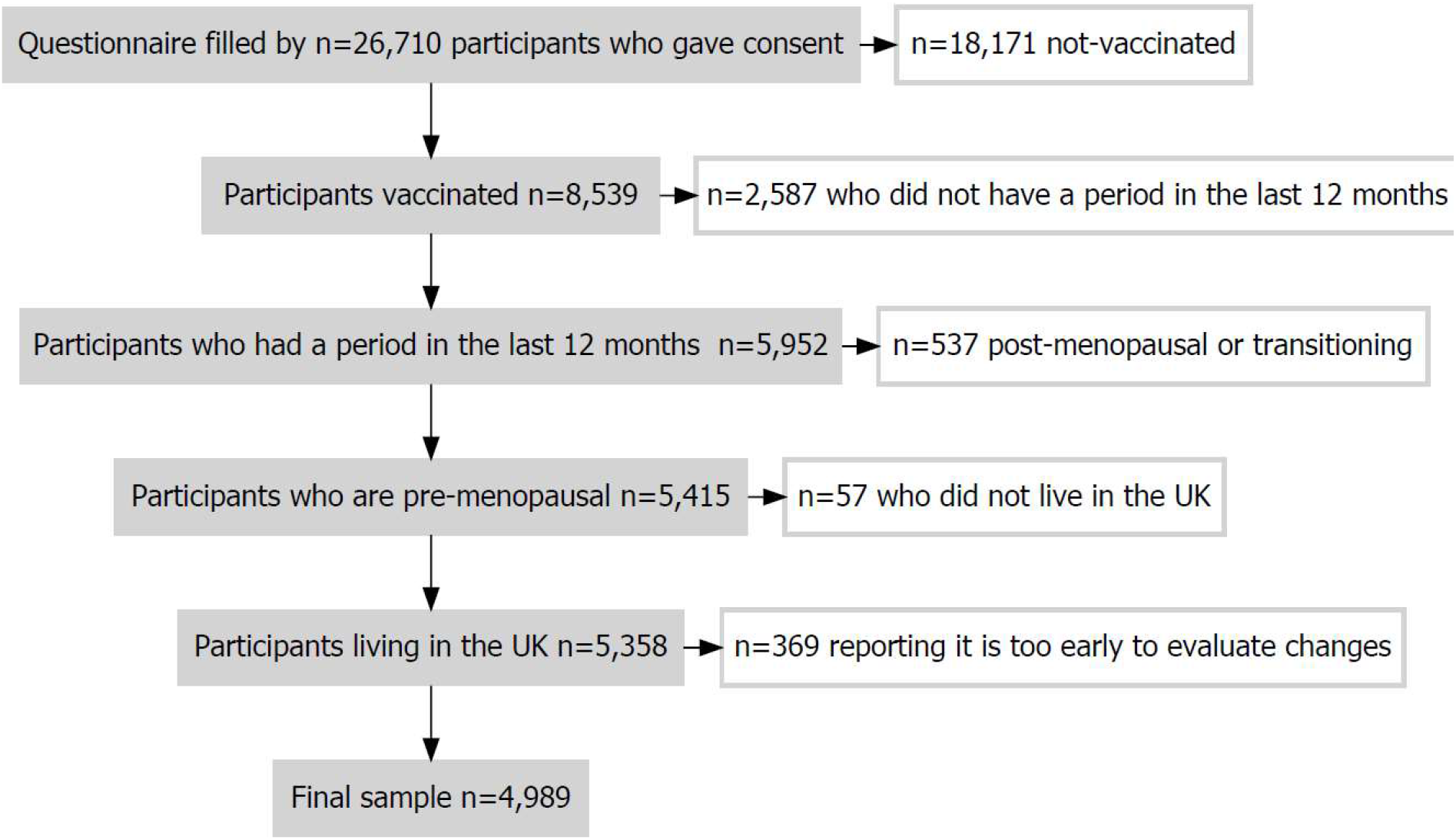
Flowchart of the study population selection.

The final sample size of vaccinated individuals is 4,989, of which 53% received the Oxford-AstraZeneca and 47% the Pfizer BioNTech vaccine (Table 1). The median age is 35 (IQR: 28 to 43) years old, with most participants living in England (81%), self-reporting as white (95%) and self-identifying as women (99%). We then grouped categories for the variables gender (women vs. other) and ethnic group (white vs. other). Although the UK vaccination campaign targeted older and at-risk populations to begin with, there does not seem to be an over-representation of over 40-year-olds. Note that 54% of participants had no deliveries and 49% had a university or college degree.

**Table 1.** Summary of the sample characteristics

| Characteristic | N = 4,989 |
| --- | --- |
| **Age, Median (IQR)** | 35 (28 – 43) |
| **Education level, n (%)** |  |
| Higher or secondary or further education (A-levels, BTEC, Baccalaureate) | 851 (17) |
| Primary & Secondary | 303 (6.2) |
| Post-graduate degree | 1,324 (27) |
| College or University | 2,395 (49) |
| Unknown | 116 |
| **Place of residence, n (%)** |  |
| UK-England | 4,031 (81) |
| UK-Northern Ireland | 159 (3.2) |
| UK-Scotland | 542 (11) |
| UK-Wales | 257 (5.2) |
| **Ethnic group, n (%)** |  |
| White | 4,734 (95) |
| Asian | 113 (2.3) |
| Black | 21 (0.4) |
| Mixed | 101 (2.0) |
| Other | 18 (0.4) |
| Unknown | 2 |
| **Net income before pandemic, n (%)** |  |
| Between £13,682 and £22,140 | 656 (15) |
| Between £22,140 and £29,254 | 614 (14) |
| Between £29,254 and £39,397 | 795 (18) |
| Between £39,397 and £76,144 | 1,453 (33) |
| Less than £13,682 | 430 (9.8) |
| More than £76,144 | 427 (9.8) |
| Unknown | 614 |
| **Smoking status before pandemic, n (%)** |  |
| I have never smoked | 3,327 (67) |
| No, but I have smoked in the past | 1,157 (23) |
| Yes, I usually smoked fewer than 10 cigarettes/day | 334 (6.7) |
| Yes, I usually smoked more than 10 cigarettes/day | 170 (3.4) |
| Unknown | 1 |
| **Marital status, n (%)** |  |
| Separated | 348 (7.2) |
| Married/partnered | 2,033 (42) |
| Nevermarried/partnered | 2,449 (50) |
| Widowed | 27 (0.6) |
| Unknown | 132 |
| **Gender, n (%)** |  |
| Man | 1 (<0.1) |
| Non Binary | 24 (0.5) |
| Other (please state) | 22 (0.4) |
| Woman | 4,939 (99) |
| Unknown | 3 |
| **Number of deliveries, n (%)** |  |
| 0 | 2,694 (54) |
| 1 | 693 (14) |
| 2 | 1,017 (20) |
| 3+ | 584 (12) |
| Unknown | 1 |
| **Contraceptive use at the time of the survey, n (%)** |  |
| Combined estradiol-progestin | 441 (11) |
| Copper IUD | 225 (5.4) |
| None | 2,421 (58) |
| Other | 84 (2.0) |
| Progestin only | 854 (21) |
| Sterilization | 130 (3.1) |
| Unknown | 834 |
| **COVID status (type), n (%)** |  |
| COVID - | 3,377 (75) |
| Long COVID | 462 (10) |
| Short COVID | 687 (15) |
| Unknown | 463 |
| **COVID status (diagnosis), n (%)** |  |
| Negative | 3,377 (76) |
| Self diagnosed + | 395 (8.9) |
| Tested + | 671 (15) |
| Unknown | 546 |
| **Number of vaccination shots, n (%)** |  |
| Yes, one shot | 4,096 (82) |
| Yes, two shots | 893 (18) |
| **Vaccine type, n (%)** |  |
| Oxford-AstraZeneca | 2,600 (53) |
| Pfizer-BioNTech | 2,335 (47) |
| Unknown | 54 |
| **Timing of 1st dose, n (%)** |  |
| Before 2021 | 331 (6.7) |
| February 2021 | 1,469 (30) |
| January 2021 | 1,497 (30) |
| March 2021 | 1,659 (33) |
| Unknown | 33 |

### Risk factors for COVID-19 vaccine-related changes in menstrual cycles

Most individuals reported no changes to their menstrual cycles following COVID-19 vaccination (80%). Only 6.1% reported more disruption, 1.5% reported less disruption and 11.5% reported “Other changes”, which, based on the previous questions participants were exposed to, could be interpreted as any changes in cycle length and regularity, period duration and volume of menstrual bleeding as well as premenstrual symptoms.

The univariable analyses show that the odds of reporting any changes to menstrual cycles after COVID-19 vaccination is associated with contraceptive type, smoking behaviour, COVID status and menstrual cycle changes over the last year (Fig. 2). All univariable models offered poor discriminative utility (AUC below 0.65, SI9). There were no differences associated with age, body mass index, ethnic group, gender, marital status, physical activity, income, education, place of residence, cycle length, period length, irregular cycles, heavy bleeding, vaccine type, vaccine timing, parity, life satisfaction changes, medication use, use of vitamins/supplements, endometriosis, PCOS, thyroid disease, uterine polyps, uterine fibroids, inter cystitis and eating disorders (Fig. 2; SI10).

**Figure 2.**
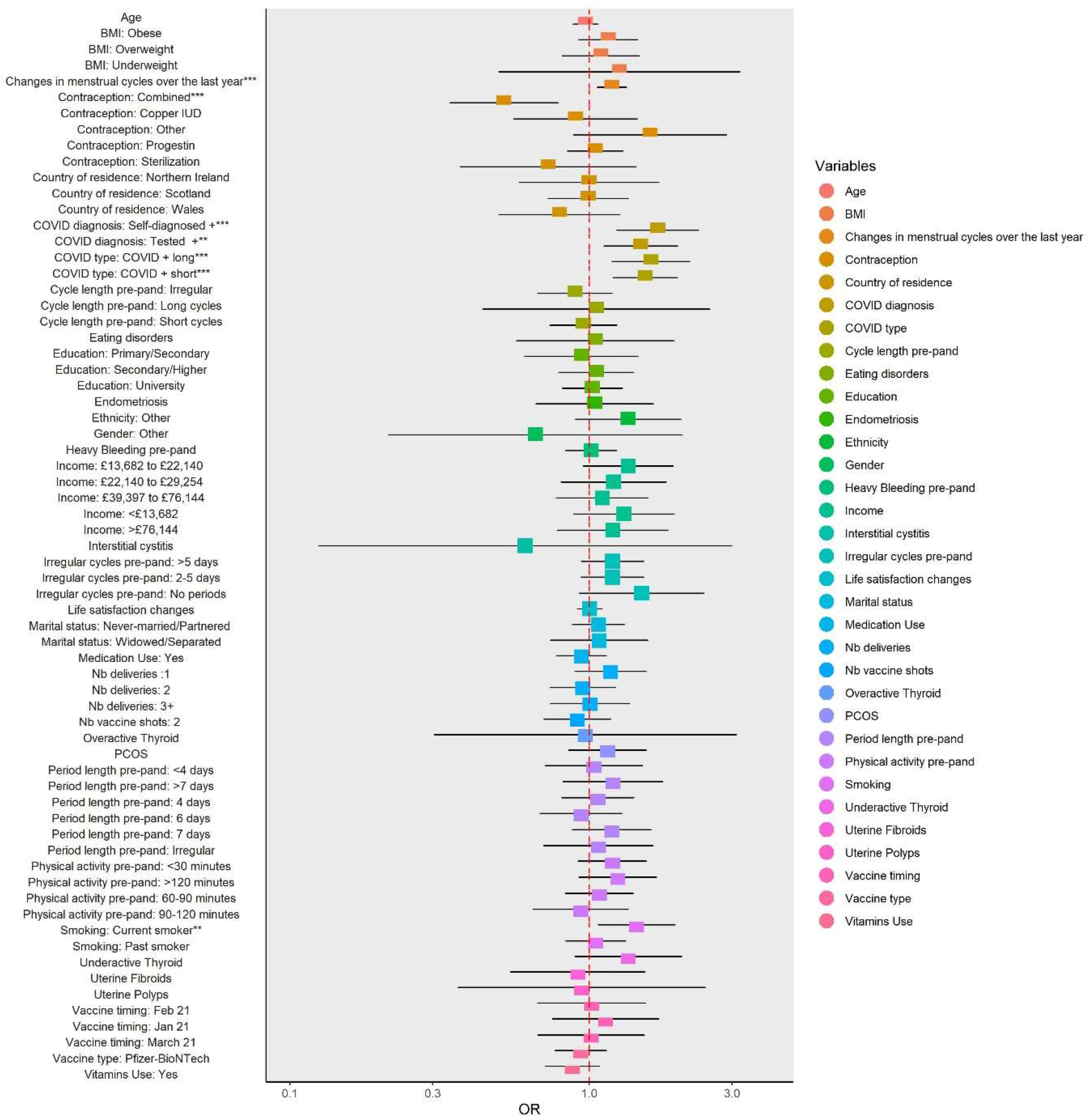
Outputs of univariable models for the odds of reporting any menstrual cycle changes following COVID-19 vaccination. The figure depicts odds-ratio and 99%CI for 33 variables. **: FDR P-value < 0.01; *** FDR P-value < 0.001.

The multivariable analyses show that the usage of combined oral contraceptives is associated with lower odds of reporting any changes by 48% (OR=0.52, 95CI=[0.34 to 0.78], *P*<0.001) while the odds of reporting any changes is increased by 44% (OR=1.44, 95CI=[1.07 to 1.94] for current smokers, *P*<0.01) and by 49 to 70% for individuals with a positive COVID status [Long Covid (OR=1.61, 95CI=[1.28 to 2.02], *P*<0.001), acute COVID (OR=1.54; 95CI=[1.27 to 1.86], *P*<0.001); self-diagnosed positive (OR=1.70, 95CI=[1.34 to 2.16], *P*<0.001), tested positive (OR=1.49, 95CI=[1.20 to 1.84], *P*<0.01), Figs 3 & 4, SI11]. The effects remain after adjusting for self-reported overall magnitude of menstrual cycle changes over the year preceding the interview (pandemic-related changes in menstrual cycle (PRCM)), which is positively associated with the risk of reporting any changes (OR=1.16, 95CI=[1.06 to 1.26], *P*<0.01). The findings were replicated when using complete cases data (SI7), indicating that the results are not an artefact of the missing data imputation process.

**Figure 3.**
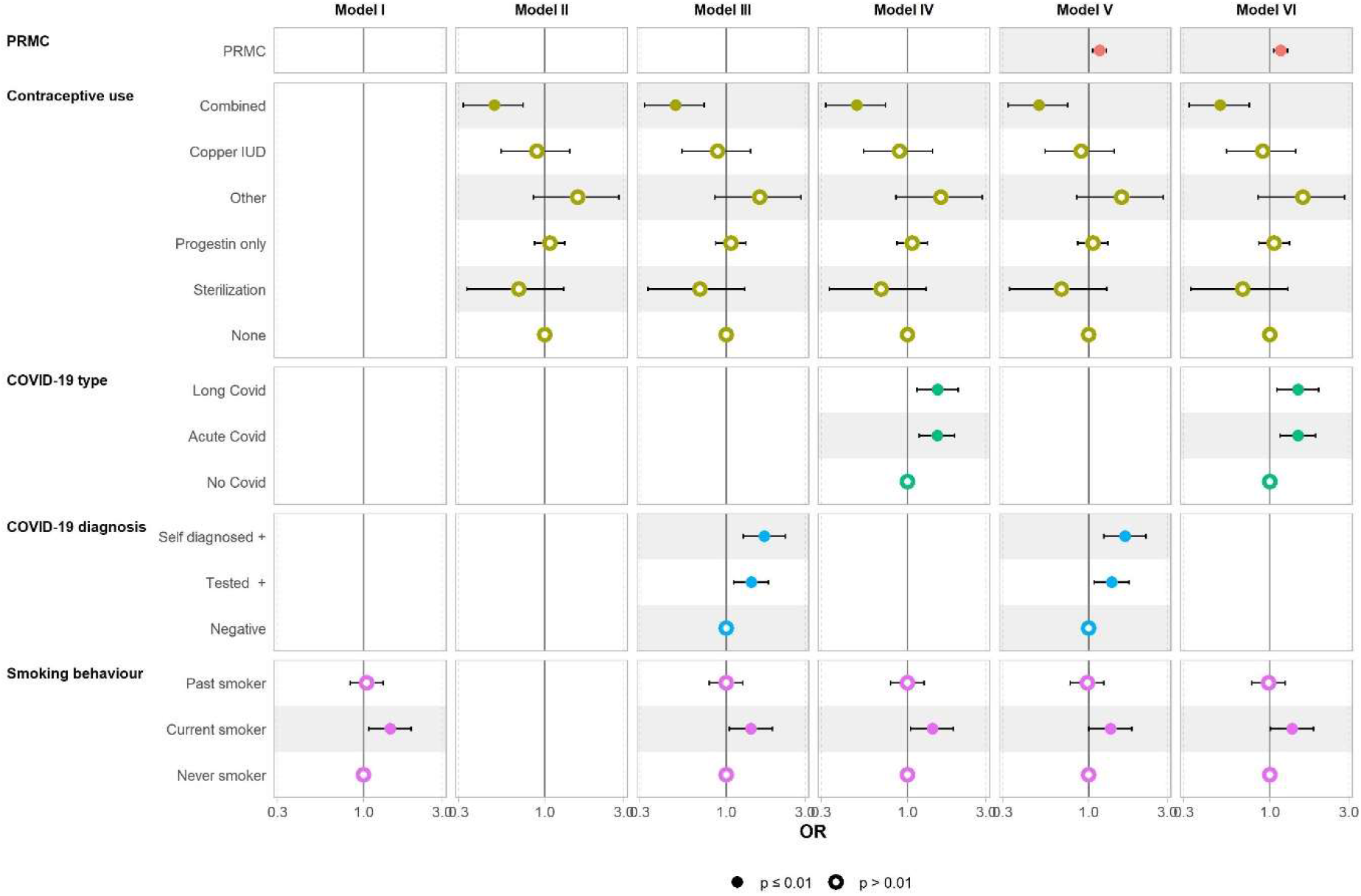
Outputs of multivariable models for the odds of reporting any menstrual cycle changes following COVID-19 vaccination. Each of the 5 exposures associated with the outcome at FDR-adjusted *P*<0.05 in the univariable analysis (i.e., pandemic-related menstrual changes (PRMC), contraceptive use, COVID-19 type, COVID-19 diagnosis, smoking behaviour) was entered in a multivariable model together with potential confounding (but not mediating) effects where appropriate (see SI5 for DAGs). *Model I*: Smoking behaviour; *Model II*: Contraceptive use; *Model III*: COVID-19 type adjusted for contraceptive use and smoking behaviour; *Model IV*: COVID-19 diagnosis adjusted for contraceptive use and smoking behaviour; *Model V*: PRMC adjusted for COVID-19 type; *Model VI*: PRMC adjusted for COVID-19 diagnosis.

**Figure 4.**
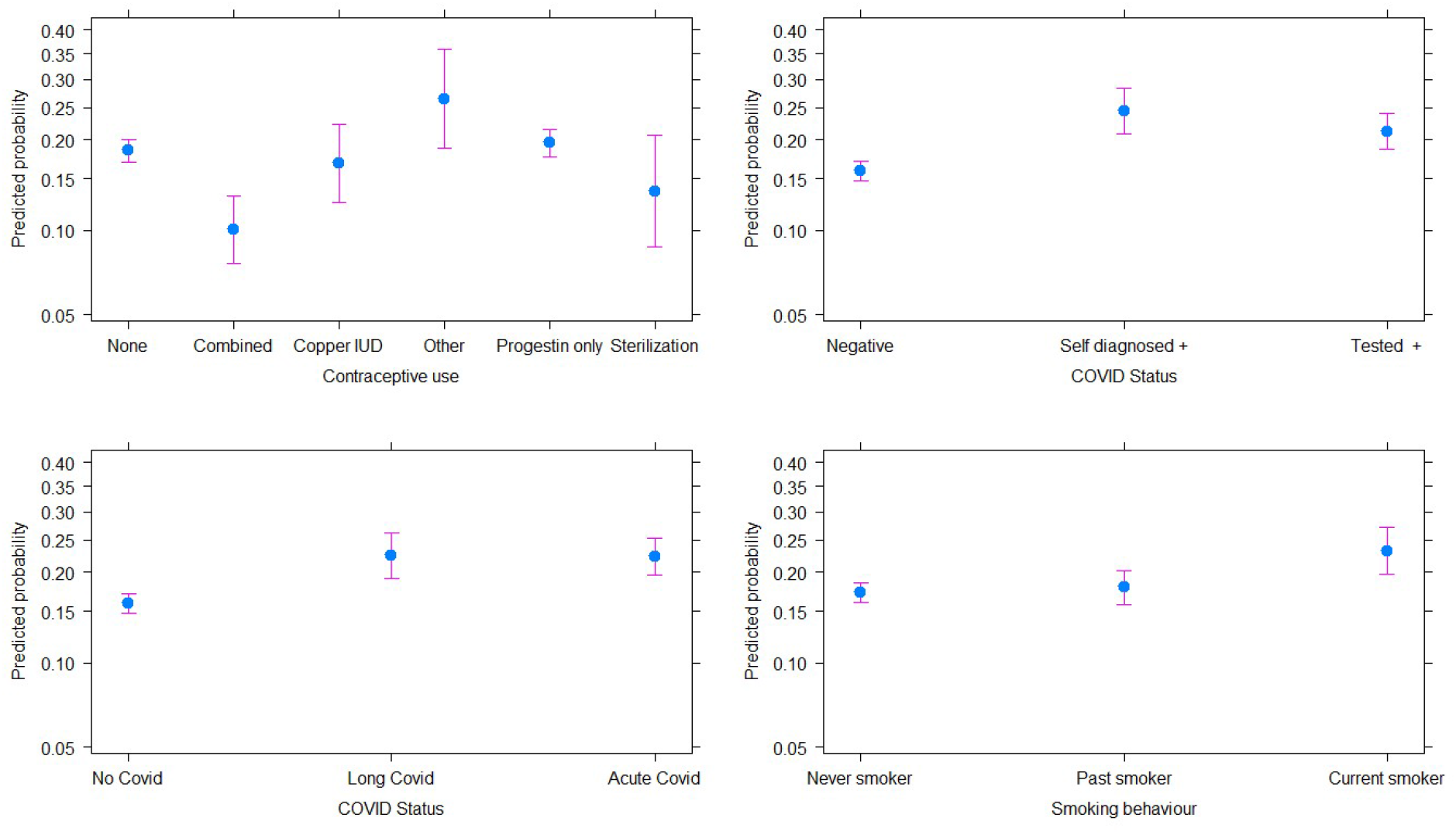
Predicted probability of reporting any menstrual changes following COVID-19 vaccination. Predicted values and 95 confidence intervals given contraceptive use, COVID status (based on type and certainty of diagnosis) and menstrual cycle changes over the last year. Most individuals (80%) reported no menstrual disturbances following COVID-19 vaccination. This probability was lower for users of combined (including oestradiol) contraceptives and higher for current smokers and those who had had a positive COVID status.

The type of contraceptive used and the history of COVID infection, while correlated, did not offer good predictive value for whether an individual will report changes to their menstrual cycle. Each exposure alone contributed an increase of only 1 to 3% of explained variance. The AUCs for the multivariate models were low across the imputed datasets (0.57 to 0.61) and the complete case dataset (0.63): the variables considered are not sufficient for predicting accurately whether an individual will report menstrual changes after vaccination. To improve the prediction accuracy of our models, we also performed a LASSO regression considering all 33 variables, but no improvement in AUC was obtained (SI12), suggesting that key variables are missing from our dataset and/or that the subjective outcome is not defined specifically enough for accurate prediction, especially if experiences of menstrual changes are diverse.

### Description of menstrual cycle changes following COVID-19 vaccination

#### Most common changes reported

The analysis of text written by participants who selected “Other changes” (n= 574, 57% of those reporting any changes) rather than “MORE disruption” or “LESS disruption” showed concerns over cycle length and menstrual bleeding patterns. The most common unigrams (individual words) were “late”, “bleed”, “early”, “long”, “heavy”, “spotting”, “short”, “pain” and “stop” and the most common bigrams (pairs of adjacent words) were “day late”, “period start”, “heavy bleed”, and “late period” (Fig. 5). While many reported menstrual cycle changes that entailed heavier bleeding/period, there was no one single pattern of symptoms, with changes including both early and late period, and diverse experiences reported (from “miss period” to “heavy bleed”).

**Figure 5.**
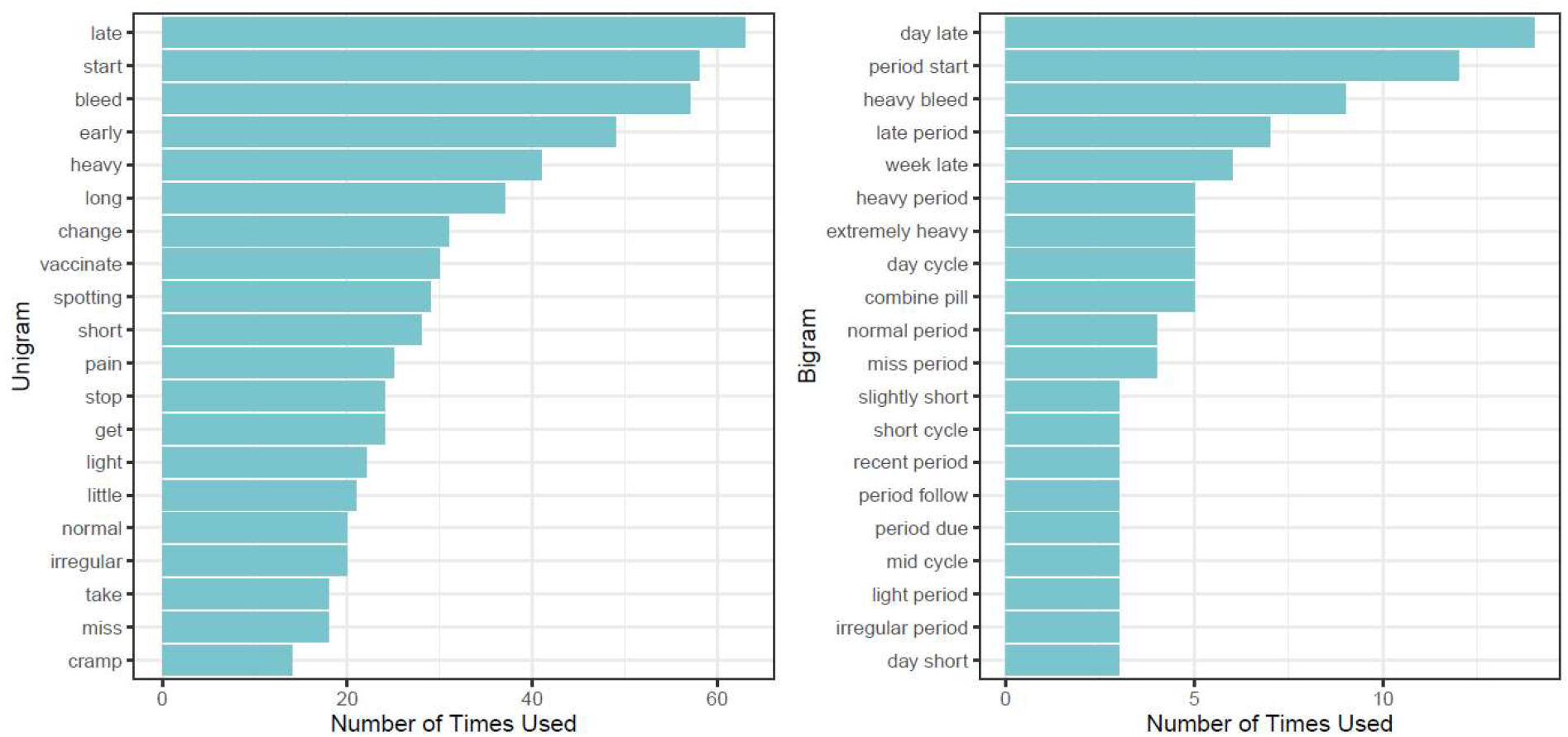
Most common words used to describe menstrual cycle changes following COVID-19 vaccination (n = 574). (A) Most common words. (B) Most common pairs of adjacent words.

#### Associations between symptoms

Only a few symptoms are correlated (φ < -0.2 or φ > 0.2). “Cramps” positively correlate with “pain” and “heavy” and “bleed” negatively correlates with “late”. Further, “lighter” positively correlates with “normal”, as participants report that “*period was two days late, and lighter than normal*”. However, “lighter” and “late” do not co-occur more than expected by chance (Fig. 6).

**Figure 6.**
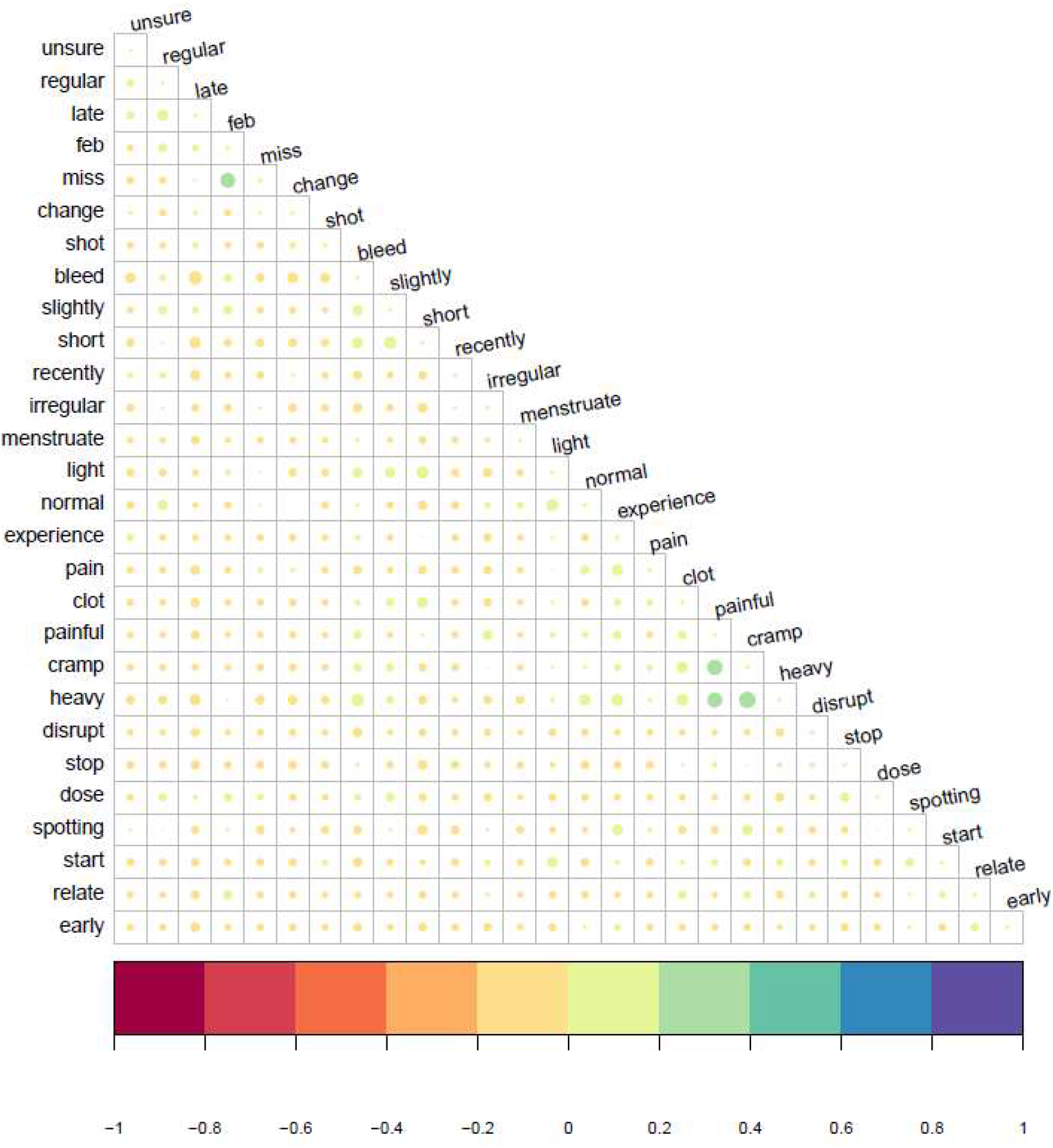
Correlation matrix between key words within sentences describing menstrual cycle changes following COVID-19 vaccination. The size and colour of the dots indicates the strength of the correlation (phi coefficient) between words.

#### Clusters of words

Different clusters of symptoms emerge from the text, such as irregular periods, heavy cramps, and pain. However, the “pain” cluster encompassed many words that are weakly correlated, suggesting a diversity of pain experience. There was also some uncertainty regarding which changes do occur, with participants finding it “*hard to say if the irregular periods are still due to covid or the vaccination*” When only correlations >0.20 were considered (Fig. 7), 4 clusters emerged: “heavy, painful, cramps”, “irregular, disruption”, “lot, clot”, and an experiential cluster “symptom, experience, pain, increase, feel”. Notably, various pain experiences that do not directly relate to menstrual cramps were reported in the main text, including stomach pain and headache.

**Figure 7.**
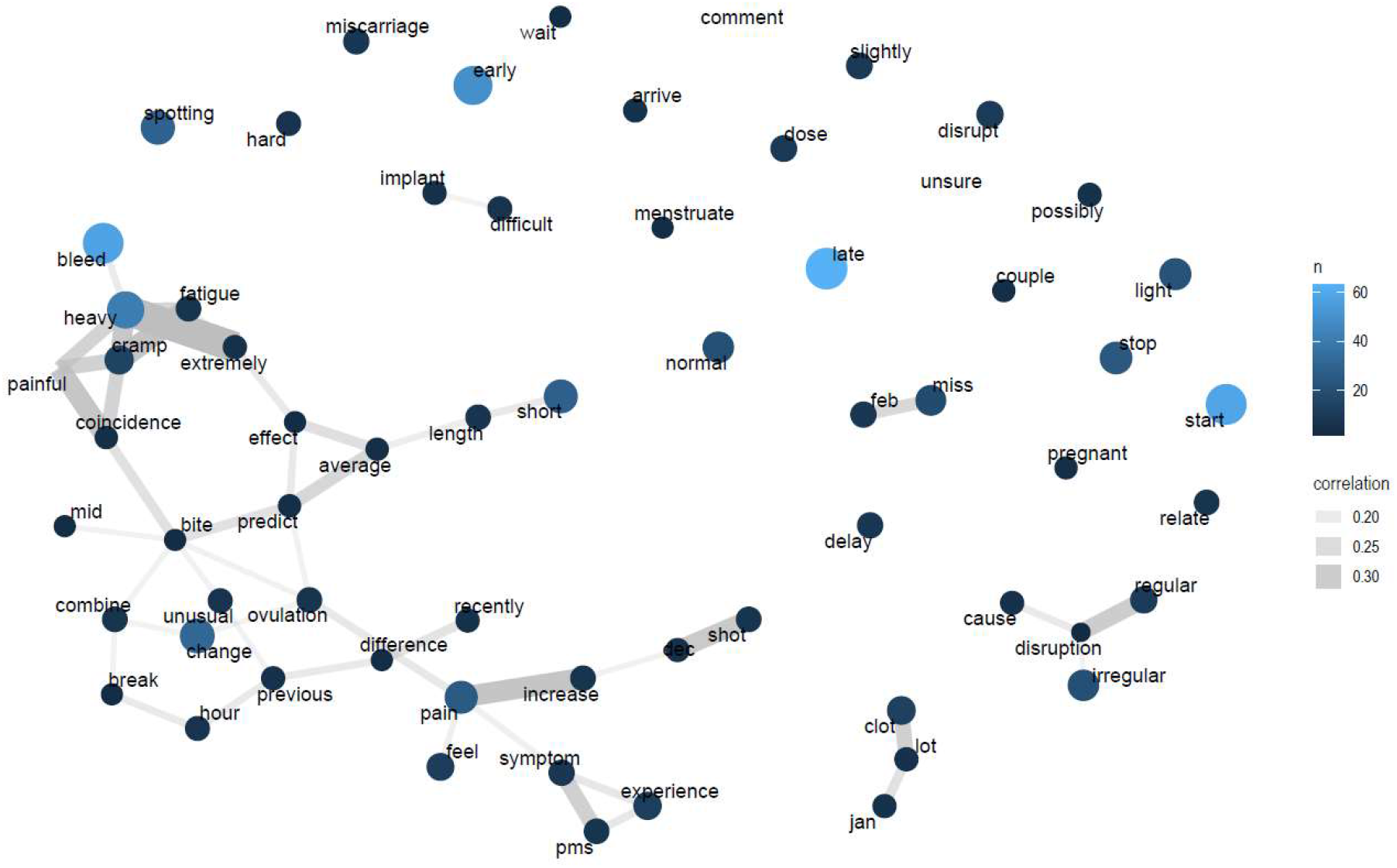
Network of words describing menstrual cycle changes following vaccination with COVID-19. Words have been lemmatised to the root of their words, for example “light” can represent both “lighter” and “light. Node size represents degree centrality (the commonality of words, only words with more than 5 occurrences are included). Edge thickness is a measure of correlation between words.

## Discussion

Using data collected in the UK prior to widespread media attention to menstrual disturbances following COVID-19 vaccination, this study found that among pre-menopausal vaccinated individuals who menstruated in the 12 months preceding the survey, 20% reported any changes to their menstrual cycles up to 4 months after receiving their first injection. In this sample, there was an association between a history of COVID infection and an increased relative risk of reporting changes of menstrual cycles following vaccination against COVID-19, independently of how COVID status was determined, i.e., using COVID type (Acute vs. Long Covid) or certainty of diagnosis (tested vs. self-diagnosed positive). This study also found that using contraceptives containing oestradiol (e.g., the pill, the vaginal ring, and the patch) is associated with a 50% lower odds of reporting menstrual cycle changes post-COVID-19 vaccination. Beyond smoking, none of the other variables investigated including age, BMI, socio-economic status, or vaccine type were associated with post-vaccination menstrual disturbances. Descriptive accounts point to diverse menstrual disturbances including “late” and “early” periods as well as “heavy bleeding” (Box 1).

### Meaning of the study

Most menstruating people in our sample did not experience menstrual changes following COVID-19 vaccination. This provides reassuring data when counselling reproductive-aged women about COVID-19 vaccination and menstrual changes. However, one in five did report menstrual disturbance following COVID-19 vaccination, a proportion that is above the threshold for a “very common” adverse reaction according to international pharmacovigilance standards. Clinicians should consider counselling women about these possible menstrual effects following COVID-19 vaccination, while emphasising the need to seek medical advice if they are severe, last more than one cycle or involve “red flag” symptoms such as inter-menstrual bleeding, post-coital bleeding, or post-menopausal bleeding. This study also suggests that current smoking and having had COVID-19 may make one more likely to experience menstrual disturbance following COVID-19 vaccination and that those on the COCP are less likely to experience menstrual disturbance. Knowledge of risk factors may help tailor advice to individuals who menstruate prior to COVID-19 vaccination.

### Strengths and weaknesses of the study

The analysis is drawing upon a survey not specifically designed to investigate the impact of COVID-19 vaccination on menstruation. It is retrospective in nature as well as sensitive to selection, recall and report biases and does not systematically assess the full spectrum of menstrual disturbance defined by the International Federation of Gynecology and Obstetrics Abnormal Uterine Bleeding System 1 [27]. We took several steps to limit selection bias during sampling (see methods) and the initial survey is broadly representative of people infected with COVID (8.9% with a positive PCR test compared to a national proportion of 6.6% at the time [28]). However, approximately 45% of the sample had received at least one dose of the vaccine, as compared to the national proportion of 59% by the time of the last survey entry [29]. In addition, menstrual changes may manifest later, and our study does not have the time depth to evaluate this possibility. However, among the studies of other vaccines conducted on a longer timescale, no effect was found by 6-9 months [14,30].

### Strengths and weaknesses of the study in relation to other studies

While the survey is also sensitive to *recall* bias, it is limited as compared to more recent surveys [8] as the issue of menstrual disturbances was not reported by the British Broadcasting Corporation until May 13, 2021 [31], as compared to a flurry of attention in US media throughout April [1–3]. Reassuringly, reporting bias would be expected to affect all sections of the sample similarly, and thus it would not explain specific associations such as with contraceptive type.

### Unanswered questions and future research

The association between a history of SARS-CoV-2 infection and menstrual disturbances post-vaccination in this study may be partly due to the effect of prior infection with SARS-CoV-2 on the immune response to vaccination, which has been found to be heightened [32]. Biological data would be needed to verify this hypothesis. The findings also suggest that exogenous oestrogen may reduce post-vaccination menstrual disturbances through anti-inflammatory or anti-viral effects. This is consistent with the recent suggestion that an “inflammatory” rather than an “ovulatory” route might explain menstrual disturbances following COVID-19 vaccination given the high prevalence of breakthrough bleeding among users of long-acting reversible contraceptives (LARC) [8]. A protective effect of oestrogen [33] and oestradiol [34] has been suggested in relation to the severity of COVID-19, and randomized control trials on unbiased samples would be needed to establish causality between oestrogen and the reduced risk of menstrual disturbances following COVID-19 vaccination. Finally, the diversity of menstrual responses to COVID-19 vaccination might be partly explained by the timing of vaccination in relation to the menstrual cycle. The findings thus call for routine menstrual data collection in COVID-19 and vaccination studies as well as research into the mechanisms of menstrual disturbance following vaccination.

## Data Availability

The anonymized data, data dictionary, scripts and SI are available on the Open Science Framework Platform (https://osf.io/pqxy2/).

**Box 1**

### What is already known on this topic?

- Menstrual disturbances including changes in frequency and/or dysmenorrhoea following vaccination have been reported as early as 1913 for the typhoid vaccine (1). Since then there have only been a few studies investigating this topic, using small sample sizes (hepatitis vaccine (2)) or reporting mixed results (HPV vaccine (3,4)).
- The UK’s Medicine and Healthcare products Regulatory Agency (MHRA) is closely monitoring reports of menstrual disorders, with more than 30,000 reports made to its yellow card surveillance scheme by 2 September 2021 following vaccination with both mRNA and adenovirus-vectored COVID-19 vaccines (5).
- In a recent preprint of a retrospective case-control study of 21,380 pre-menopausal participants living in the US, 45.8% of 9,579 people with regular menstrual cycles experienced heavier bleeding after COVID-19 vaccination. In addition, 70.5% of 1,545 non-menstruating people using long-acting reversible contraceptives (LARC) experienced breakthrough bleeding after COVID-19 vaccination (6). This informative study may be affected by selection bias and may not be generalisable.

### What this study adds

- In a large sample of participants vaccinated against COVID-19 surveyed in the UK before widespread media attention to related menstrual changes, the prevalence of menstrual changes was 1 in 5.
- Out of 33 socio-demographic, health, vaccine, COVID- and pandemic-related and reproductive variables, the odds of reporting any menstrual changes following COVID-19 vaccination were associated with a history of SARS-CoV-2 infection, smoking behaviour and the type of contraceptives used.
- Menstrual changes that were reported were diverse, ranging from increased bleeding to the cessation of bleeding.
- The study highlights the need for greater consideration of the menstrual cycle in health interventions.

## Supporting Information Caption

SI1: Recruitment facebook ads

SI2 : Information sheet

SI3 : Survey questions

SI4 : Operationalization of variables

SI5 : DAG

SI6: Missing data evaluation

SI7: Complete cases analysis

SI8 : Text analysis

SI9 : AUC univariable models

SI10: Table univariable models

SI11 : Table multivariable models

SI12: AUC Lasso

## Notes

### Competing Interest Statement

The authors have declared no competing interest.

### Funding Statement

The British Academy

### Author Declarations

The study has been reviewed by, and received ethics clearance through, the Oxford University School of Anthropology and Museum Ethnography Departmental Research Ethics Committee [SAME_C1A_20_029]

### Summary of Updates

Typo in abstract

